# Maternal late-pregnancy serum unmetabolized folic acid concentrations are not associated with infant allergic disease - A prospective cohort study

**DOI:** 10.1101/2020.10.29.20222588

**Authors:** Karen P Best, Tim J Green, Dian Sulistyoningrum, Thomas R Sullivan, Susanne Aufreiter, Susan L Prescott, Maria Makrides, Monika Skubisz, Deborah L O’Connor, Debra J Palmer

**Affiliations:** Women and Kids Theme, South Australian Health and Medical Research Institute, Adelaide, Australia; Adelaide Medical School, Faculty of Health and Medical Sciences, The University of Adelaide, Australia; School of Public Health, University of Adelaide, Australia; Department of Nutritional Sciences, Faculty of Medicine, University of Toronto, Toronto, Ontario, Canada; Translational Medicine, Research Institute, Hospital for Sick Children, Toronto, Ontario, Canada; The ORIGINS Project, Telethon Kids, Nedlands, WA, Australia; Department of Immunology, Perth Children’s Hospital, Nedlands, WA, Australia; Telethon Kids Institute, The University of Western Australia, Nedlands, WA, Australia; School of Medicine, The University of Western Australia, Crawley, WA, Australia

**Keywords:** unmetabolized folic acid, allergic disease, atopic dermatitis, eczema, food allergy, folate, folic acid, infant, pregnancy

## Abstract

**Background:** The increase in childhood allergic disease in recent decades has coincided with increased folic acid intakes during pregnancy. Circulating unmetabolized folic acid (UMFA) has been proposed as a biomarker of excessive folic acid intake.

**Objective:** We aimed to determine if late-pregnancy serum UMFA and total folate concentrations were associated with allergic disease risk in the offspring at one year of age in a population at high risk of allergy.

**Methods:** The cohort consisted of 561 mother-infant pairs from Western Australia. To be eligible the infant had a first-degree relative (mother, father or sibling) with a history of medically diagnosed allergic disease. Maternal serum was collected between 36 and 40 weeks of gestation. UMFA concentrations were measured by tandem mass spectrometry using stable isotope dilution, folate concentrations were determined using the microbiological method with standardized kits. Infant allergic disease outcomes of medically diagnosed eczema, steroid treated eczema, atopic eczema, IgE-mediated food allergy, allergen sensitization and medically diagnosed wheeze were assessed at 1 year of age.

**Results:** Median (IQR) for UMFA and serum folate was 1.6 (0.6-4.7) and 53.2 (32.6-74.5) nmol/L, respectively. Of the infants, 34.6% had medically diagnosed eczema, 26.4% allergen sensitization and 14.9% had an IgE-mediated food allergy. In both adjusted and unadjusted models there was little evidence of association between UMFA or serum folate and any of the infant allergy outcomes.

**Conclusion:** In this cohort of children at high risk for allergic disease there was no association between maternal UMFA or serum folate measured in late pregnancy and allergic disease outcomes at 1 year of age.

## Introduction

The prevalence of early childhood atopic diseases, predominately atopic dermatitis (eczema) and food allergy, have increased over recent decades. There is emerging evidence that immune function at birth is predictive of whether a child will develop allergic disease (1-4). Therefore, the in-utero period may be critical in determining immune development trajectories towards an allergic phenotype (1, 5) and maternal diet during pregnancy could be a potential modifiable early life influence of allergic disease development.

This increase in early life allergic diseases has coincided with an increase in folic acid intake during the perinatal period. Women are advised to take folic acid containing supplements (6, 7) prior to and during early pregnancy to reduce the incidence of neural tube defects (NTDs). (8-10) In addition, to further reduce NTDs, more than 80 countries worldwide have mandated the addition of folic acid to food staples such as flour. (11) NTDs occur in the first month of pregnancy, yet many women continue to take folic acid-containing supplements throughout pregnancy with no known benefit.

Folic acid is a synthetic form of folate; due to its high bioavailability, stability and low cost, it is used in supplements and for food fortification. Once consumed, folic acid is normally converted to tetrahydrofolate, by the enzyme dihydrofolate reductase (DHFR), before being further converted to 5-methyltetrahydrofolate. Higher intakes of folic acid can saturate the capacity of DHFR leading to the presence of unmetabolized folic acid (UMFA) in circulation. (29-32). Circulating UMFA has been detected in pregnant women and in cord blood (33-36). There is speculation that UMFA in circulation is a biomarker of excessive folic acid intake and may be causing harm through epigenetic changes to fetal gene expression, with subsequent increased disease risk (37, 38).

Animal models have shown that pregnant mice fed diets high in folic acid exhibit altered expression of immune genes through changes in DNA methylation in the offspring. Such changes have been associated with enhanced severity of allergic airway disease. (12)

Several observational cohort studies have reported inconsistent associations between higher prenatal folic acid or folate intakes and risk of allergic disease in the offspring (13-16) (17-27) however many rely on dietary assessment to measure exposure. Of the studies that examined biomarkers to determine exposure, only one differentiated between specific forms of folate (20). This nested case control study from the United States, reported that UMFA concentrations in cord blood were associated with an increased risk of food allergy, but not food allergen sensitization, however other allergic disease outcomes were not reported. (21)

Australia is an ideal setting to examine associations between folic acid in pregnancy and allergic disease outcomes in the offspring. It has among the highest prevalence of allergic disorders in the developed world in addition to high prenatal folic acid exposures from food fortification and high rates of prenatal folic acid supplementation. (28, 29) To date, no studies have examined the association between maternal late gestation UMFA and infant allergic disease. We aimed to determine if maternal serum UMFA or folate concentrations in late pregnancy predicted infant allergic disease outcomes at one year of age, in a pregnancy cohort of women carrying a fetus at high hereditary risk of allergic disease (history of allergic disease in at least one immediate family member).

## Methods

### Study Population

The data presented here come from mother-infant pairs from a prospective cohort study in Perth, Western Australia. The study was designed to explore whether maternal diet, lifestyle and environmental factors influence offspring susceptibility to allergic disease. Pregnant women >18 years of age whose unborn infant had a first-degree relative (mother, father or sibling) with a history of medically diagnosed allergic disease (asthma, allergic rhinitis, eczema and/or food allergy) were recruited from local participating maternity antenatal clinics and classes between November 2011 and December 2016. Women were >36 weeks gestation at enrolment, with a singleton pregnancy, non-smokers (while pregnant) and healthy with no known complications (including immunodeficiency, pre-eclampsia, and major congenital anomalies). The original cohort study was approved by the Princess Margaret Hospital Human Research Ethics Committee (1942EP), and all participants provided written informed consent. The maternal blood analysis for this study was also approved by the Women’s and Children’s Health Network Human Research Ethics Committee in 2019 (HREC/19/WCHN/21).

The investigators in the original cohort aimed to have around 600 mother–infant pairs with infant allergy outcomes at one year of age. This was based on previous cohorts which have examined associations between maternal diet in pregnancy and allergic disease outcomes in infants with a high hereditary risk of allergic disease. (18, 30)

### Maternal Data and Blood Collection

Maternal baseline data were collected between 36 and 40-weeks’ gestation, including history of allergic disease, education, ethnicity, parity, and pet ownership (cat, dog, or both).

Maternal non-fasting blood samples were collected from the cubital vein into a serum clot activator tube (Vacuette, Z Serum Clot activator; Greiner Bio-One GmbH, Kremsmünster, Austria). The blood samples were allowed to clot, spun at 4000rpm for 10 minutes, serum was aliquoted into 1-2ml tubes and stored at ^-^80°C until analyzed.

### Infant Allergic Disease Assessment and Definitions

At one year of age the participating infants were assessed at the Princess Margaret Hospital in Perth, Australia. A parent was asked if the infant ever had eczema which was diagnosed by a medical doctor (medically diagnosed eczema) during the first year of life, and if the eczema skin lesions were responsive to topical steroid treatment prescribed by a medical doctor (steroid treated eczema). The parent was also asked if the infant had any wheeze symptoms which had been diagnosed by a medical doctor (medically diagnosed wheeze). Immunoglobulin-E (IgE) -mediated food allergy was based on history of immediate IgE-mediated symptoms (within 60 minutes of food ingestion) including angioedema, urticaria, cough, wheeze, stridor, vomiting, diarrhea, cardiovascular symptoms, and allergen sensitization to the same food detected by positive skin prick test (SPT) at the one year of age visit. SPT was conducted to detect allergen sensitization to common Australian food and environmental allergens including cow’s milk, hen’s egg, peanut, cashew nut, wheat, rye grass, house dust mite, and cat (Hollister-Stier Laboratories, Spokane, WA, USA), as well as histamine as a positive control. A response was considered positive if the mean of the horizontal and perpendicular wheal diameter was 3 mm or greater in size than the mean wheal of the negative control site at 15 minutes. Sensitization was defined as a positive skin prick test result to at least one of the allergens assessed. Each infant’s SPT and clinical allergy assessment results were confirmed by the research physician. Infant birth details (including delivery mode, gestational weight, gestational age and infant sex) were also collected.

### Serum Unmetabolized Folic Acid (UMFA)

Between August 2019 and July 2020 maternal serum samples were analyzed for UMFA by stable isotope dilution-Tandem mass spectrometry following the methods of Pfeiffer et.al. (31) using spectrometrically verified standards of folic acid and an internal standard of 13C5-folic acid (Merck, Switzerland). Briefly, samples and standard spiked with folic acid-[13C5] in 1 % ammonium formate, 0.5% ascorbic acid buffer (pH 3.2) were loaded onto phenyl cartridges (1mL) (Phenomenex, Torrance. CA) previously conditioned with 2 mL each of methanol, acetonitrile and 1 % ammonium formate buffer (pH 3.2) and were allowed to equilibrate for 1 minute. The loaded cartridges were washed sequentially with 3mL of 0.05 % ammonium formate buffer (pH=3.4, 0.25% ascorbic acid), and folic acid was eluted from the columns using an elution buffer (0.5mL) of 49% water, 40% methanol, 10% acetonitrile, 1% concentrated acetic acid and 0.5% ascorbic acid. Eluted samples were stored at −80 CC until analysis by tandem mass spectrometry at The Analytical Facility for Bioactive Molecules, The Hospital for Sick Children, Toronto, Canada.

Eluted solutions were separated chromatographically on a Kinetex PFP (50 x 3.0 mm, 2.6 µm particle size) column (Phenomenex, Torrance CA). The mass-to-charge ratios of the transitions of interest, m/z 442.4 → m/z 295.1 for folic acid and m/z 447.4 → 495.1 for 13C folic acid, were monitored using an AB Sciex QTRAP 5500 triple quadrupole MS system (Agilent 1290 UHPLC system, (Agilent Technologies, Santa Clara, CA, USA

The inter-batch accuracy and precision were determined with the use of NIST 1950 Standard Reference Material with a certified value of 1.51± 0.45 ng/mL. Each group of samples was analyzed along with an aliquot of the reference material NIST1950, Metabolites in Frozen Human Plasma; the mean (±SD) obtained for 12 batches of samples was 1.80± 0.21 ng/mL, with a CV of 11.9%. Some UMFA concentrations were below the limit of detection and were set to the mid-point between 0 and the detection limit for analysis. and were set to the mid-point between 0 and the detection limit for analysis.

### Serum folate

Serum folate concentrations were determined using the microbiological method based on the technique of O’Broin and Kelleher, using standardized kits from the US Centers for Disease Control and Prevention (Atlanta, GA).[18–20] This method uses 96 well microplates, 5-methyl tetrahydrofolate (Merck) as the calibrator, and chloramphenicol resistant Lactobacillus rhamnosus (ATCC® 27773^™^) as the test organism. High- and low-quality controls for serum folate provided by the Centers for Disease Control were run in quadruplets on every plate.

### Statistical Analysis

Associations between the maternal folate measures and infant allergy outcomes were evaluated using logistic regression, with effects described as odds ratios with 95% confidence intervals. UMFA and serum folate concentrations were treated as continuous exposures in the main analysis, with the assumption of a linear association with the log odds of each allergic disease outcome assessed using Hosmer-Lemeshow tests. For completeness, additional analyses were also performed with the UMFA and folate measures grouped into quartiles and treated as categorical exposures. For each outcome variable and UMFA and folate measure, both unadjusted and adjusted analyses were performed, with adjustment for maternal age, further maternal education after high school, maternal Caucasian ethnicity, maternal cat/dog ownership, maternal parity > 1, vaginal delivery, infant sex, infant birth weight, and infant gestational age at birth. (32) UMFA concentrations below the limit of detection were set to the mid-point between 0 and the detection limit for analysis. All statistical analyses were performed using Stata version 16.0 (College Station, TX: StataCorp LP).

## Results

### Study Population

561 mother–infant pairs with complete maternal data, maternal blood sample and at least one of the infant allergic disease outcome measures were included in this analysis. Maternal and infant characteristics for the mother–infant pairs are summarized in **Table 1**. The majority of the participating women were European Caucasian (91%) and most had completed post-secondary school education (75%). All infants had at least one immediate family member (first-degree relative) with a history of allergic disease and 92% had maternal allergic disease.

**Table 1.** Descriptive statistics for maternal and infant characteristics.

| **Characteristic** | **Total N=561** |
| --- | --- |
| Maternal age (years) | 32.9 (4.8) |
| Maternal allergic disease | 517/560 (92.3%) |
| Maternal positive skin prick test | 482/537 (89.8%) |
| Maternal atopy | 467/560 (83.4%) |
| Maternal European Caucasian ethnicity | 493/544 (90.6%) |
| Further maternal education after secondary school | 417/558 (74.7%) |
| Maternal dog and/or cat ownership | 334/559 (59.7%) |
| Maternal parity > 1 | 296/560 (52.9%) |
| Maternal late pregnancy vitamin use | 457/528 (86.6%) |
| Maternal dietary folate intake at 32-36 weeks gestation (mg/day) | 265 (212-329) |
| Vaginal delivery | 372/550 (67.6%) |
| Infant GA at birth (weeks) [n=550] | 39.3 (1.1) |
| Infant birth weight (grams) [n=559] | 3502 (434) |
| Female infant sex | 271/561 (48.3%) |

Data are presented as mean (SD) or median (IQR) for continuous measures, and n/total (%) for categorical measures. GA, gestational age.

**Table 2.** Associations between unmetabolized folic acid concentration (continuous) and infant allergic disease outcomes^1^

| **Outcome** | **Unadjusted odds ratio (95% CI)** | **P-value** | **Adjusted<sup>2</sup> odds ratio (95% CI)** | **Adjusted p-value** |
| --- | --- | --- | --- | --- |
| Medically diagnosed eczema | 0.99 (0.89, 1.09) | 0.77 | 1.02 (0.90, 1.15) | 0.76 |
| Steroid treated eczema | 1.02 (0.93, 1.13) | 0.66 | 1.04 (0.92, 1.18) | 0.54 |
| Atopic eczema | 1.05 (0.94, 1.17) | 0.39 | 1.06 (0.92, 1.22) | 0.44 |
| IgE mediated food allergy | 1.04 (0.93, 1.16) | 0.52 | 1.05 (0.91, 1.21) | 0.52 |
| Allergen sensitization | 1.02 (0.92, 1.13) | 0.74 | 1.02 (0.90, 1.16) | 0.77 |
| Medically diagnosed wheeze | 1.08 (0.96, 1.21) | 0.22 | 1.13 (0.96, 1.34) | 0.14 |

<sup>1</sup> Odds ratios describe the effect of a 10 nmol/L increase in unmetabolized folic acid; goodness of fit confirmed using Hosmer Lemeshow tests
<sup>2</sup> Adjusted for maternal age, further maternal education after high school, maternal Caucasian ethnicity, infant sex, infant birth weight, infant GA at birth, maternal cat/dog ownership, parity > 1 and vaginal delivery

### Maternal serum unmetabolized folic acid and folate concentrations

In late gestation, UMFA was detectable in 520/559 (93.0%) of maternal serum samples, with concentrations ranging from 0.03 to 244.7 (median 1.6; IQR 0.6 to 4.7) nmol/L. Maternal serum folate concentration ranged from 4.3 to 185.0 (median 53.2; IQR 32.6 to74.5) nmol/L. The Spearman rank correlation coefficient between the UMFA and serum folate concentrations was 0.50 (95% CI 0.44 to 0.57).

### Infant allergic disease outcomes

Of the infants, 194/561 (34.6%) had medically diagnosed eczema and 150/561 (26.7%) had eczema requiring steroid treatment during the first year of life. The allergen sensitization rate was 26.4% (146/552), with 14.9% (83/558) of infants classified as having atopic eczema (medically diagnosed eczema and allergen sensitization) and 14.9% (83/558) of infants with an IgE-mediated food allergy. Only 8.7% (49/561) infants had medically diagnosed wheeze during infancy.

No associations were found between maternal UMFA (**Table 3**) or folate concentrations (**Table 3**) and infant allergic disease outcomes. In the case of UMFA, results weren’t sensitive to the method used to deal with values below the detection limit (set to the limit, set to 0 or excluded from analysis, results didn’t change). Similarly, we did not find any associations between maternal UMFA quartiles (**Table 4**) or folate quartiles (**Table 5**) and any of the infant allergic disease outcomes in additional adjusted and unadjusted (not shown) analyses.

**Table 3.** Associations between serum folate (continuous) and infant allergic disease outcomes^1^.

| **Outcome** | **Unadjusted odds ratio (95% CI)** | **P-value** | **Adjusted<sup>2</sup> odds ratio (95% CI)** | **Adjusted p-value** |
| --- | --- | --- | --- | --- |
| Medically diagnosed eczema | 0.99 (0.94, 1.05) | 0.81 | 1.00 (0.95, 1.06) | 0.93 |
| Steroid treated eczema | 1.01 (0.96, 1.07) | 0.69 | 1.01 (0.95, 1.07) | 0.76 |
| Atopic eczema | 1.04 (0.97, 1.11) | 0.27 | 1.03 (0.96, 1.10) | 0.43 |
| IgE mediated food allergy | 0.95 (0.88, 1.02) | 0.16 | 0.93 (0.86, 1.00) | 0.07 |
| Allergen sensitization | 1.00 (0.95, 1.06) | 0.87 | 0.99 (0.94, 1.05) | 0.78 |
| Medically diagnosed wheeze | 1.02 (0.94, 1.11) | 0.57 | 1.03 (0.95, 1.12) | 0.51 |

<sup>1</sup> Odds ratios describe the effect of a 10 nmol/L increase in serum folate; goodness of fit confirmed using Hosmer Lemeshow tests
<sup>2</sup> Adjusted for maternal age, further maternal education after high school, maternal Caucasian ethnicity, infant sex, infant birth weight, infant GA at birth, maternal cat/dog ownership, parity > 1 and vaginal delivery

**Table 4.**
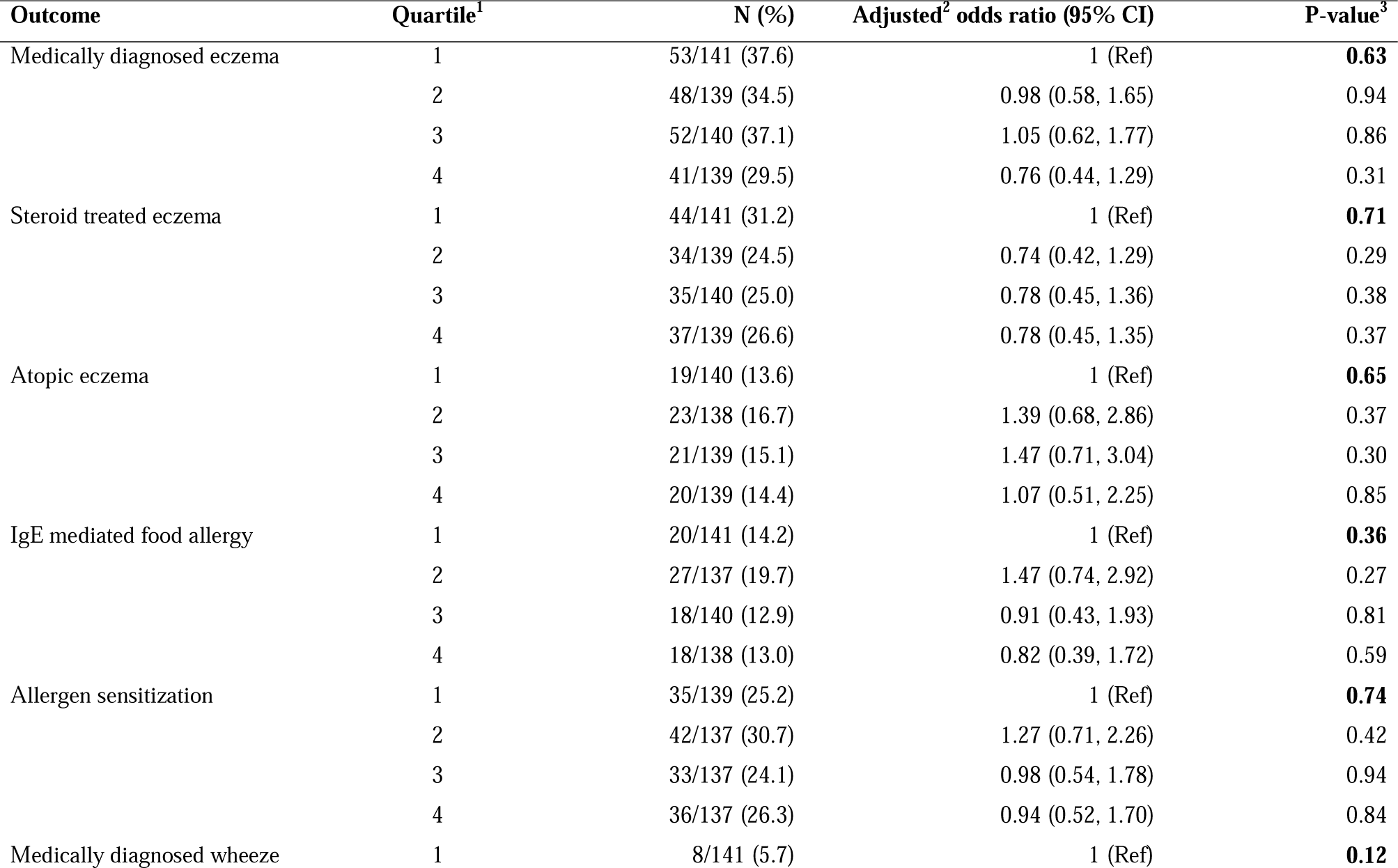
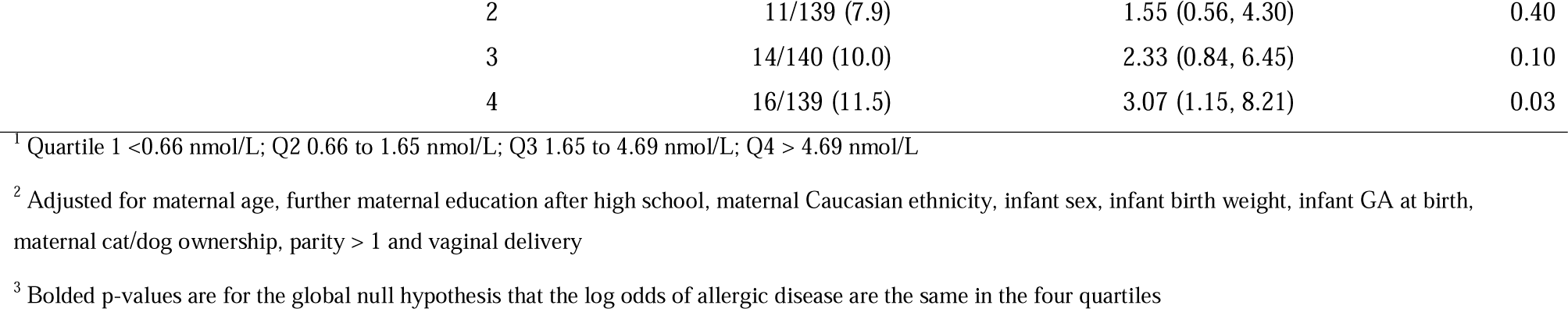
Associations between unmetabolized folic acid concentration (quartiles) and infant allergic disease outcomes

| **Outcome** | **Quartile<sup>1</sup>** | **N (%)** | **Adjusted<sup>2</sup> odds ratio (95% CI)** | **P-value<sup>3</sup>** |
| --- | --- | --- | --- | --- |
| Medically diagnosed eczema | 1 | 53/141 (37.6) | 1 (Ref) | **0.63** |
|  | 2 | 48/139 (34.5) | 0.98 (0.58, 1.65) | 0.94 |
|  | 3 | 52/140 (37.1) | 1.05 (0.62, 1.77) | 0.86 |
|  | 4 | 41/139 (29.5) | 0.76 (0.44, 1.29) | 0.31 |
| Steroid treated eczema | 1 | 44/141 (31.2) | 1 (Ref) | **0.71** |
|  | 2 | 34/139 (24.5) | 0.74 (0.42, 1.29) | 0.29 |
|  | 3 | 35/140 (25.0) | 0.78 (0.45, 1.36) | 0.38 |
|  | 4 | 37/139 (26.6) | 0.78 (0.45, 1.35) | 0.37 |
| Atopic eczema | 1 | 19/140 (13.6) | 1 (Ref) | **0.65** |
|  | 2 | 23/138 (16.7) | 1.39 (0.68, 2.86) | 0.37 |
|  | 3 | 21/139 (15.1) | 1.47 (0.71, 3.04) | 0.30 |
|  | 4 | 20/139 (14.4) | 1.07 (0.51, 2.25) | 0.85 |
| IgE mediated food allergy | 1 | 20/141 (14.2) | 1 (Ref) | **0.36** |
|  | 2 | 27/137 (19.7) | 1.47 (0.74, 2.92) | 0.27 |
|  | 3 | 18/140 (12.9) | 0.91 (0.43, 1.93) | 0.81 |
|  | 4 | 18/138 (13.0) | 0.82 (0.39, 1.72) | 0.59 |
| Allergen sensitization | 1 | 35/139 (25.2) | 1 (Ref) | **0.74** |
|  | 2 | 42/137 (30.7) | 1.27 (0.71, 2.26) | 0.42 |
|  | 3 | 33/137 (24.1) | 0.98 (0.54, 1.78) | 0.94 |
|  | 4 | 36/137 (26.3) | 0.94 (0.52, 1.70) | 0.84 |
| Medically diagnosed wheeze | 1 | 8/141 (5.7) | 1 (Ref) | **0.12** |

|  |  |  |  |
| --- | --- | --- | --- |
| 2 | 11/139 (7.9) | 1.55 (0.56, 4.30) | 0.40 |
| 3 | 14/140 (10.0) | 2.33 (0.84, 6.45) | 0.10 |
| 4 | 16/139 (11.5) | 3.07 (1.15, 8.21) | 0.03 |

<sup>1</sup> Quartile 1 <0.66 nmol/L; Q2 0.66 to 1.65 nmol/L; Q3 1.65 to 4.69 nmol/L; Q4 > 4.69 nmol/L
<sup>2</sup> Adjusted for maternal age, further maternal education after high school, maternal Caucasian ethnicity, infant sex, infant birth weight, infant GA at birth, maternal cat/dog ownership, parity > 1 and vaginal delivery
<sup>3</sup> Bolded p-values are for the global null hypothesis that the log odds of allergic disease are the same in the four quartiles

**Table 5.**
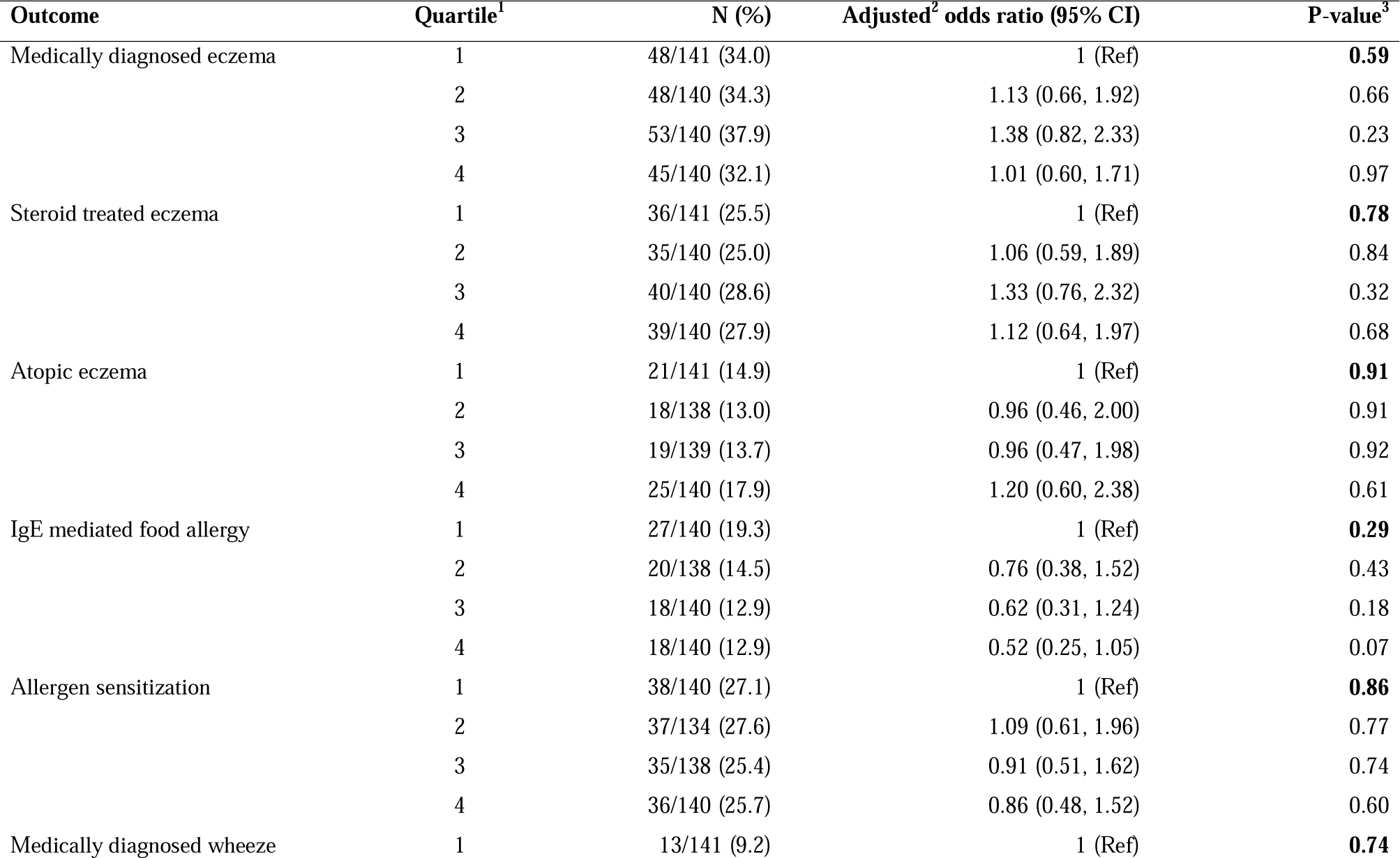
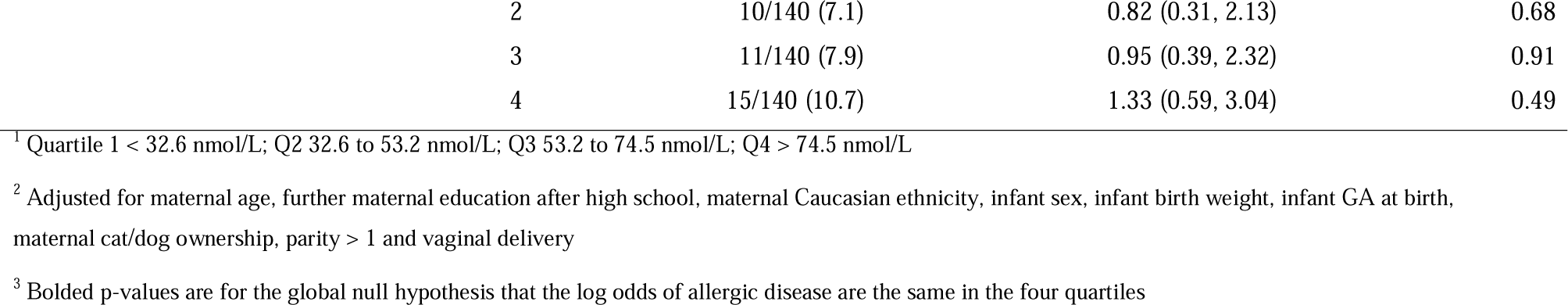
Associations between serum folate (quartiles) and infant allergic disease outcomes.

| **Outcome** | **Quartile<sup>1</sup>** | **N (%)** | **Adjusted<sup>2</sup> odds ratio (95% CI)** | **P-value<sup>3</sup>** |
| --- | --- | --- | --- | --- |
| Medically diagnosed eczema | 1 | 48/141 (34.0) | 1 (Ref) | **0.59** |
|  | 2 | 48/140 (34.3) | 1.13 (0.66, 1.92) | 0.66 |
|  | 3 | 53/140 (37.9) | 1.38 (0.82, 2.33) | 0.23 |
|  | 4 | 45/140 (32.1) | 1.01 (0.60, 1.71) | 0.97 |
| Steroid treated eczema | 1 | 36/141 (25.5) | 1 (Ref) | **0.78** |
|  | 2 | 35/140 (25.0) | 1.06 (0.59, 1.89) | 0.84 |
|  | 3 | 40/140 (28.6) | 1.33 (0.76, 2.32) | 0.32 |
|  | 4 | 39/140 (27.9) | 1.12 (0.64, 1.97) | 0.68 |
| Atopic eczema | 1 | 21/141 (14.9) | 1 (Ref) | **0.91** |
|  | 2 | 18/138 (13.0) | 0.96 (0.46, 2.00) | 0.91 |
|  | 3 | 19/139 (13.7) | 0.96 (0.47, 1.98) | 0.92 |
|  | 4 | 25/140 (17.9) | 1.20 (0.60, 2.38) | 0.61 |
| IgE mediated food allergy | 1 | 27/140 (19.3) | 1 (Ref) | **0.29** |
|  | 2 | 20/138 (14.5) | 0.76 (0.38, 1.52) | 0.43 |
|  | 3 | 18/140 (12.9) | 0.62 (0.31, 1.24) | 0.18 |
|  | 4 | 18/140 (12.9) | 0.52 (0.25, 1.05) | 0.07 |
| Allergen sensitization | 1 | 38/140 (27.1) | 1 (Ref) | **0.86** |
|  | 2 | 37/134 (27.6) | 1.09 (0.61, 1.96) | 0.77 |
|  | 3 | 35/138 (25.4) | 0.91 (0.51, 1.62) | 0.74 |
|  | 4 | 36/140 (25.7) | 0.86 (0.48, 1.52) | 0.60 |
| Medically diagnosed wheeze | 1 | 13/141 (9.2) | 1 (Ref) | **0.74** |

|  |  |  |  |
| --- | --- | --- | --- |
| 2 | 10/140 (7.1) | 0.82 (0.31, 2.13) | 0.68 |
| 3 | 11/140 (7.9) | 0.95 (0.39, 2.32) | 0.91 |
| 4 | 15/140 (10.7) | 1.33 (0.59, 3.04) | 0.49 |

<sup>1</sup> Quartile 1 < 32.6 nmol/L; Q2 32.6 to 53.2 nmol/L; Q3 53.2 to 74.5 nmol/L; Q4 > 74.5 nmol/L
<sup>2</sup> Adjusted for maternal age, further maternal education after high school, maternal Caucasian ethnicity, infant sex, infant birth weight, infant GA at birth, maternal cat/dog ownership, parity > 1 and vaginal delivery
<sup>3</sup> Bolded p-values are for the global null hypothesis that the log odds of allergic disease are the same in the four quartiles

## Discussion

This is the first prospective cohort study to examine the association between maternal late pregnancy UMFA status and multiple allergic disease outcomes in a ‘high-risk’ infant population. We found no evidence of associations between maternal UMFA or folate concentrations and any infant allergic disease outcomes.

One other cohort study has examined the association between UMFA and childhood allergy outcomes using cord blood to measure exposure. (20) In contrast to our findings, McGowan et. al. reported that children whose cord blood concentrations were in the highest quartile of UMFA (n=14/33) had an 8.5-fold (95% CI 1.7 to 42.8) increased risk of confirmed food allergy compared to the lowest quartile. (20) However, interestingly there was no association between UMFA and food allergen sensitization. Furthermore, atopic dermatitis/eczema outcomes (the most common allergic disease in infancy) were not reported. Given there was only 6.6% (33/502 children with cord blood UMFA) of the children with confirmed food allergy in the McGowan et. al. publication, and the wide confidence interval, there is a possibility of this being a chance finding. Our Australian based study differed from this US study in several ways. Notably, our cohort had higher rates of offspring food allergy (14.9% vs 6.6%). In addition, our population included mostly women of European Caucasian ethnicity (91%), whereas, the Boston Birth Cohort in the study by McGowan et. al. were predominately of non-Hispanic black ethnicity (only 7% ‘white’). We cannot compare our UMFA concentrations in maternal serum to those reported in cord blood samples in the study by McGowan et al. However, the prevalence and range of detectable concentrations of UMFA in our maternal study population (93%, range: undetectable to 245nmol/L) are similar to those reported by Plumptre et. al. from blood samples collected from women in early pregnancy (56% Caucasian) and taking a folic acid containing supplement (97%; range undetectable to 244 nmol/L). (33)

We also found no association between maternal serum folate at 36-40 weeks’ gestation and infant allergic disease outcomes even though we did find higher median maternal serum folate concentrations (53.2 (IQR 32.6-74.5) nmol/L), compared to a previous Western Australian cohort study (n=435), where average serum folate concentrations were 37.2 nmol/I (IQR 25.6–50.5 nmol/I) after 28 weeks gestation. (18) The previous Western Australian cohort study also did not find any associations between maternal serum folate concentrations and infant eczema outcomes, but did find that cord blood folate concentrations <50 nmol/l (OR = 3.02; 95% CI 1.16–7.87) and >75 nmol/l (OR = 3.59; 95% CI 1.40–9.20) were associated with greater infant allergen sensitization risk than cord blood folate concentrations between 50 and 75 nmol/l. (18)

Previous findings from observational studies examining associations between folate exposure in pregnancy from food and/or supplement use and offspring allergic disease have been equivocal, the majority reporting an increased risk, (13-16) (17-23); some no association, (24, 25) and others suggesting a protective effect of folate (26, 27). Two studies suggested a “U shaped distribution” association where both high and low folate exposure increased allergy risk (19, 33), however we could not find any similar associations in our study. The inconsistency in results may be due to heterogeneity of populations studied including genetic differences and background folate intakes from food. Study design differences including; timing and classification (i.e. diet, supplement use, or folate biomarkers) of folate/folic acid exposure, as well as offspring age and types of allergic disease outcomes assessed may also confound results. Many of the observational studies showing associations between folic acid and allergic disease have relied on maternal reported folic acid supplementation or dietary assessment methods as the exposure, which can be subject to reporting bias and measurement error. Furthermore, folic acid supplements are usually consumed as part of a multi-vitamin and mineral supplement and hence it is impossible to determine the independent effect of folicacid. In our study there was no association between late pregnancy supplement use and risk of allergic disease despite 86% of participants taking folic acid containing supplements.

A strength of our study is the objective measurement of serum UMFA and folate concentrations which reduce potential reporting bias due to reliance on participant memory of dietary intake of folate from foods or supplement use. However, the pregnant women in our study were not fasted and the timing of the maternal blood sample collection in relation to the last ingestion of maternal folic acid supplementation is unknown. UMFA rises rapidly after ingesting folic acid and falls steadily over time. Our cohort of infants at high hereditary risk of allergic disease increased our incidence of allergy outcomes compared to studies in a general population. However, we cannot exclude the fact that these infants with a genetic tendency towards atopy may have a predetermined disposition to an allergic phenotype which may not be modifiable by maternal folic acid concentration.

We controlled for a number of important confounders including those known to be associated with allergy risk, but as with any epidemiological study, the possibility of confounding remains. As an example, as with the incidence of allergic disease (34), higher folic acid supplement use has also been associated with higher socioeconomic status. Moreover, as folic acid is usually taken as part of a multi-vitamin and mineral preparation, other nutrients also may influence allergy outcomes both positive and negative.

In conclusion, we found no associations between maternal late gestation serum UMFA or folate levels and risk of infant allergic disease at one year of age in a population with high hereditary risk of atopy. Further work, including randomized controlled trials with objective biomarkers are needed to confirm that high folic acid exposure during late pregnancy, largely driven by the combination of food fortification and prenatal folic acid supplement use, does not increase the risk of childhood allergic disease in the general population.

## Data Availability

Data will be available to Researchers who provide a methodologically sound research proposal following review and approval by the trial steering committee and completion of a signed data access agreement.
Following approval, de‐identified, individual participant data that underlie the results reported in this article (text, tables, figures and appendices) and/or dataset(s) will be limited to those participants and variables that are necessary for completion of the approved research proposal.
Data sharing requests for de‐identified raw data should be made to the trial steering committee and can be submitted to

## Abbreviations

UMFA: unmetabolized folic acid
NTDs: neural tube defects
DHFR: dihydrofolate reductase
IgE: Immunoglobulin-E
SPT: skin prick test

## Acknowledgements

We would like to acknowledge the women and children who participated in the cohort study, and associated research team staff who have made this work possible.

## Authors’ contributions to the manuscript

KPB, TJG, DJP, MM, MS and TRS designed this research project. KPB provided day to day study oversight; DS and SA conducted analysis of samples with oversight from TS and DLO. SLP and DJP provided maternal blood samples and maternal and infant data, including the allergic disease outcome data. TRS performed statistical analysis; KPB drafted the manuscript with input from TJG and DJP. All authors reviewed the manuscript for final content.

## References

1. Tulic MK, Hodder M, Forsberg A, McCarthy S, Richman T, D’Vaz N, van den Biggelaar AH, Thornton CA, Prescott SL. Differences in innate immune function between allergic and nonallergic children: new insights into immune ontogeny. Journal of Allergy Clinical Immunology 2011;127(2):470-8. e1.

2. Osborne NJ, Koplin JJ, Martin PE, Gurrin LC, Lowe AJ, Matheson MC, Ponsonby A-L, Wake M, Tang ML, Dharmage SCJJoA. Prevalence of challenge-proven IgE-mediated food allergy using population-based sampling and predetermined challenge criteria in infants. J Journal of Allergy Clinical Immunology 2011;127(3):668-76. e2.

3. Prescott S, Allen KJ. Food allergy: riding the second wave of the allergy epidemic. Pediatric allergy and immunology : official publication of the European Society of Pediatric Allergy and Immunology 2011;22(2):155–60. doi: 10.1111/j.1399-3038.2011.01145.x.

4. Mullins RJ, Dear KB, Tang ML. Time trends in Australian hospital anaphylaxis admissions in 1998-1999 to 2011-2012. J Allergy Clin Immunol 2015;136(2):367–75. doi: 10.1016/j.jaci.2015.05.009.

5. Pfefferle PI, Buchele G, Blumer N, Roponen M, Ege MJ, Krauss-Etschmann S, Genuneit J, Hyvarinen A, Hirvonen MR, Lauener R, et al. Cord blood cytokines are modulated by maternal farming activities and consumption of farm dairy products during pregnancy: the PASTURE Study. J Allergy Clin Immunol 2010;125(1):108-15 e1-3. doi: 10.1016/j.jaci.2009.09.019.

6. Australia GoS. South Australian Perinatal Practice Guidelines - Vitamin and mineral supplementation in pregnancy. In: Ageing DfHa, ed.: SA Maternal & Neonatal Clinical Network 2015.

7. Houk VN, Oakley GP, Erickson JD, Mulinare J, James LM. Recommendations for the use of folic acid to reduce the number of cases of spina bifida and other neural tube defects. 1992.

8. Group MVSR. Prevention of neural tube defects: results of the Medical Research Council Vitamin Study. MRC Vitamin Study Research Group. Lancet 1991;338(8760):131–7. doi: 10.1016/0140-6736(91)90133-a.

9. Berry RJ, Li Z, Erickson JD, Li S, Moore CA, Wang H, Mulinare J, Zhao P, Wong LY, Gindler J, et al. Prevention of neural-tube defects with folic acid in China. China-U.S. Collaborative Project for Neural Tube Defect Prevention. N Engl J Med 1999;341(20):1485–90. doi: 10.1056/NEJM199911113412001.

10. Czeizel AE, Dudas I. Prevention of the first occurrence of neural-tube defects by periconceptional vitamin supplementation. N Engl J Med 1992;327(26):1832–5. doi: 10.1056/NEJM199212243272602.

11. Initiative FF. Global Progress of Industrially Milled Cereal Grains. 2019.

12. Hollingsworth JW, Maruoka S, Boon K, Garantziotis S, Li Z, Tomfohr J, Bailey N, Potts EN, Whitehead G, Brass DM, et al. In utero supplementation with methyl donors enhances allergic airway disease in mice. J Clin Invest 2008;118(10):3462–9. doi: 10.1172/JCI34378.

13. Yang L, Jiang L, Bi M, Jia X, Wang Y, He C, Yao Y, Wang J, Wang Z. High dose of maternal folic acid supplementation is associated to infant asthma. Food Chem Toxicol 2015;75:88–93. doi: 10.1016/j.fct.2014.11.006.

14. Bekkers MB, Elstgeest LE, Scholtens S, Haveman-Nies A, de Jongste JC, Kerkhof M, Koppelman GH, Gehring U, Smit HA, Wijga AH. Maternal use of folic acid supplements during pregnancy, and childhood respiratory health and atopy. European Respiratory Journal 2012;39(6):1468–74.

15. Whitrow MJ, Moore VM, Rumbold AR, Davies MJ. Effect of supplemental folic acid in pregnancy on childhood asthma: a prospective birth cohort study. Am J Epidemiol 2009;170(12):1486–93. doi: 10.1093/aje/kwp315.

16. Haberg SE, London SJ, Stigum H, Nafstad P, Nystad W. Folic acid supplements in pregnancy and early childhood respiratory health. Arch Dis Child 2009;94(3):180–4. doi: 10.1136/adc.2008.142448.

17. Kiefte-de Jong JC, Timmermans S, Jaddoe VW, Hofman A, Tiemeier H, Steegers EA, de Jongste JC, Moll HA. High Circulating Folate and Vitamin B-12 Concentrations in Women During Pregnancy Are Associated with Increased Prevalence of Atopic Dermatitis in Their Offspring. The Journal of nutrition 2012;142(4):731–8.

18. Dunstan JA, West C, McCarthy S, Metcalfe J, Meldrum S, Oddy WH, Tulic MK, D’Vaz N, Prescott SL. The relationship between maternal folate status in pregnancy, cord blood folate levels, and allergic outcomes in early childhood. Allergy 2012;67(1):50–7. doi: 10.1111/j.1398-9995.2011.02714.x.

19. Parr CL, Magnus MC, Karlstad O, Haugen M, Refsum H, Ueland PM, McCann A, Nafstad P, Haberg SE, Nystad W, et al. Maternal Folate Intake during Pregnancy and Childhood Asthma in a Population-based Cohort. American journal of respiratory and critical care medicine 2017;195(2):221–8. doi: 10.1164/rccm.201604-0788OC.

20. McGowan EC, Hong X, Selhub J, Paul L, Wood RA, Matsui EC, Keet CA, Wang X. Association Between Folate Metabolites and the Development of Food Allergy in Children. The journal of allergy and clinical immunology In practice 2020;8(1):132–40 e5. doi: 10.1016/j.jaip.2019.06.017.

21. Veeranki SP, Gebretsadik T, Mitchel EF, Tylavsky FA, Hartert TV, Cooper WO, Dupont WD, Dorris SL, Hartman TJ, Carroll KN. Maternal Folic Acid Supplementation During Pregnancy and Early Childhood Asthma. Epidemiology 2015;26(6):934–41. doi: 10.1097/EDE.0000000000000380.

22. Liu J, Li Z, Ye R, Liu J, Ren A. Periconceptional folic acid supplementation and risk of parent-reported asthma in children at 4–6 years of age. J ERJ open research 2020;6(1).

23. Zetstra-van der Woude PA, De Walle HE, Hoek A, Bos HJ, Boezen HM, Koppelman GH, de Jong-van den Berg LT, Scholtens S. Maternal high-dose folic acid during pregnancy and asthma medication in the offspring. Pharmacoepidemiol Drug Saf 2014;23(10):1059–65. doi: 10.1002/pds.3652.

24. Molloy AM, Kirke PN, Brody LC, Scott JM, Mills JL. Effects of folate and vitamin B12 deficiencies during pregnancy on fetal, infant, and child development. Food Nutr Bull 2008;29(2 Suppl):S101-11; discussion S12-5. doi: 10.1177/15648265080292S114.

25. Granell R, Heron J, Lewis S, Davey Smith G, Sterne JA, Henderson J. The association between mother and child MTHFR C677T polymorphisms, dietary folate intake and childhood atopy in a population-based, longitudinal birth cohort. Clinical and experimental allergy : journal of the British Society for Allergy and Clinical Immunology 2008;38(2):320–8. doi: 10.1111/j.1365-2222.2007.02902.x.

26. Fortes C, Mastroeni S, Mannooranparampil TJ, Di Lallo D. Pre-natal folic acid and iron supplementation and atopic dermatitis in the first 6 years of life. Arch Dermatol Res 2019;311(5):361–7. doi: 10.1007/s00403-019-01911-2.

27. Kim JH, Jeong KS, Ha EH, Park H, Ha M, Hong YC, Bhang SY, Lee SJ, Lee KY, Lee SH, et al. Relationship between prenatal and postnatal exposures to folate and risks of allergic and respiratory diseases in early childhood. Pediatr Pulmonol 2015;50(2):155–63. doi: 10.1002/ppul.23025.

28. Zhou SJ, Best K, Gibson R, McPhee A, Yelland L, Quinlivan J, Makrides M. Study protocol for a randomised controlled trial evaluating the effect of prenatal omega-3 LCPUFA supplementation to reduce the incidence of preterm birth: the ORIP trial. BMJ Open 2017;7(9):e018360. doi: 10.1136/bmjopen-2017-018360.

29. Valera-Gran D, Garcia de la Hera M, Navarrete-Munoz EM, Fernandez-Somoano A, Tardon A, Julvez J, Forns J, Lertxundi N, Ibarluzea JM, Murcia M, et al. Folic acid supplements during pregnancy and child psychomotor development after the first year of life. JAMA Pediatr 2014;168(11):e142611. doi: 10.1001/jamapediatrics.2014.2611.

30. West CE, Dunstan J, McCarthy S, Metcalfe J, D’Vaz N, Meldrum S, Oddy WH, Tulic MK, Prescott SL. Associations between maternal antioxidant intakes in pregnancy and infant allergic outcomes. Nutrients 2012;4(11):1747–58.

31. Pfeiffer CM, Fazili Z, McCoy L, Zhang M, Gunter EW. Determination of folate vitamers in human serum by stable-isotope-dilution tandem mass spectrometry and comparison with radioassay and microbiologic assay. Clin Chem 2004;50(2):423–32. doi: 10.1373/clinchem.2003.026955.

32. Nurmatov U, Nwaru BI, Devereux G, Sheikh A. Confounding and effect modification in studies of diet and childhood asthma and allergies. Allergy 2012;67(8):1041–59. doi: 10.1111/j.1398-9995.2012.02858.x.

33. Plumptre L, Masih SP, Ly A, Aufreiter S, Sohn KJ, Croxford R, Lausman AY, Berger H, O’Connor DL, Kim YI. High concentrations of folate and unmetabolized folic acid in a cohort of pregnant Canadian women and umbilical cord blood. Am J Clin Nutr 2015;102(4):848–57. doi: 10.3945/ajcn.115.110783.

34. Mullins RJ, Clark S, Camargo CA, Jr. Socio-economic status, geographic remoteness and childhood food allergy and anaphylaxis in Australia. Clinical and experimental allergy : journal of the British Society for Allergy and Clinical Immunology 2010;40(10):1523–32. doi: 10.1111/j.1365-2222.2010.03573.x.

